# Assessing Sociodemographic Factors Associated with Household Hardships during the COVID-19 Pandemic in Manhiça, Mozambique using Data Collected between April 2021 and February 2022

**DOI:** 10.1101/2023.12.21.23300355

**Authors:** Ariel Q. Nhacolo, Jonathan A. Muir, Zachary J. Madewell, Fatima Keiri, Charfudin N. Sacoor, Edgar L. Jamisse, Elisio G. Xerinda, Teodimiro Matsena, Aura M. Hunguana, Quique Bassat, Cynthia G. Whitney, Inácio M. Mandomando, Solveig A. Cunningham

## Abstract

**Background:** COVID-19 resulted in vast disruption to life in the 21^st^ century. To quell the disease spread, national governments implemented several containment measures like state of emergency, curfews, and lockdowns that likely created hardships for households. To improve knowledge of the negative consequences of these lockdowns, we examine the extent to which the pandemic period was associated with hardships at the household level and assess factors associated with household vulnerability to these hardships.

**Methods:** We conducted a cross-sectional survey between April 2021 and February 2022 among households residing in the district of Manhiça through a survey questionnaire fielded in the Health and Demographic Surveillance System (HDSS) operating in Manhiça, Mozambique. Logistic regression was used to analyze associations between the head of household and household characteristics with specific household hardships (business closure, food price increase, household member detained, input inflation, job loss).

**Results:** Households headed by individuals with lower education and employed in non-agricultural occupations as well as households that were larger in size or poorer in asset ownership compared to other households were generally at greater risk of experiencing a larger variety of hardships. Conversely, households that owned “distance demolishing technologies” such as motorcycles were less likely to experience these hardships, presumably as they were able to transcend local conditions.

**Conclusions:** These results identify at-risk groups according to a social determinants of vulnerability framework and will help inform future policies and practices that aim to mitigate the negative consequences of COVID-19 as well as future disease outbreaks.

## Background

Beyond direct health effects,[1] the COVID-19 pandemic resulted in vast disruption to life. As government-enforced containment measures such as curfews and lockdowns were implemented to stem the pace and severity of the disease, a variety of social and economic consequences ensued.[2] To adhere to these lockdowns, many businesses temporarily closed their in-person workplaces; while for some, these closures were permanent. Mitigation efforts also affected healthcare providers, who had to restrict in-person access to patients and/or limit services. These efforts may have affected already vulnerable populations by limiting access to healthcare and food.[3-15] These indirect effects, in turn, would have exacerbated the direct effects of SARS-CoV-2 by increasing the risk of undernutrition and other conditions that heighten the risk of serious illness.[6, 15] Population-based knowledge of the extent to which households living in resource-limited countries have experienced resource restrictions and other hardships has remained limited due to limitations in population-based surveillance.[16]

In sub-Saharan Africa (SSA), national governments undertook considerable efforts to quell the spread of COVID-19.[2, 17] In Mozambique, the first confirmed COVID-19 case was detected on Mar 22, 2020. On Mar 31, 2020, the Mozambique government declared a state of emergency to slow the spread of the pandemic, followed by a gradual reopening of the economy.[18, 19] The state of emergency included the closure of schools, suspension of entertainment and sports events, limited hours of marketing, reduced public transportation occupancy, and limiting in-person presence at workplaces to one-third of the workforce.[19] These measures resulted in decreased travel and movement and limited public services and gatherings.[20] Early implementation of COVID-19 prevention measures in Mozambique kept COVID-19 cases relatively low in the country from March to September 2020.[18, 21] However, these efforts likely affected vulnerable populations in myriad ways beyond direct effects on disease transmission.

Vulnerability is the communal effect of cultural, economic, political, and social processes that alter the experience of and recovery from a given hazard.[22] Disasters, particularly in the context of environmental hazards, are often not the result of a given hazard, but rather of the shock to individual and community coping patterns and the inputs and outputs of social systems.[23-25] Social vulnerability is partially the result of social disparities that shape or influence the susceptibility of different groups to hazards while also controlling their capacity to respond.[26, 27] Individual and household level factors often associated with vulnerability include demographic characteristics such as age, ethnicity, race, and sex; socioeconomic status (e.g., lower wealth, employment, and/or education); household composition (e.g., presence of children); and housing and transportation.[28] Social vulnerability also involves place disparities stemming from characteristics of the built environment; e.g., differential availability of resources between urban and rural areas may exacerbate individual- and household-level vulnerabilities.[29-32] To understand the broader consequences of the pandemic, it is important to consider economic, political, and social markers of vulnerability.

Mozambique is one of the poorest countries in the world.[33] Following decades of wars (liberation war during 1964-1974, and civil war during 1976-1992) the poverty level in Mozambique declined from 70% to about 50% between 1996 and 2008.[18] However, this economic recovery flattened and deteriorated in 2015 and the following years, mainly due to the interruption of funding from key donors.[33, 34] Other reasons for the economic stagnation included severe weather shocks (e.g., droughts, cyclones, and flooding), increased violence in the central and northern regions of the country, and the international economic crisis.[33, 35] These conditions resulted in substantial population displacement, destruction of infrastructure, and increased risk of infectious disease.[36] These shocks further exacerbated economic disparities within the country. Southern and urban areas experienced less economic impact compared to rural areas. Also, despite the decline in the poverty rates following the war in Mozambique, high population growth led to an increase in the number of individuals living in poverty.[18] In Mozambique, the pandemic hit an already vulnerable population.

Mozambique’s economy lost 5.6 percentage points of growth in the second quarter of 2020 due to COVID-19 and the state of emergency.[37] This economic contraction was mainly due to reduced exports, reflecting the dependence on a limited range of export commodities, rendering it susceptible to trade-related shocks. Limited marketing for agriculture caused by transport restrictions, closed borders, and reduced purchasing power forced farmers to lower their prices, contributing to an agricultural shock.[38] In places where most of the population were self-employed or employees for wages, particularly in urban areas, the impacts were higher because there was no national social welfare system to support these people. These factors increased overall vulnerability to economic hardship and food insecurity during the pandemic.[39] However, the effects of the pandemic varied according to socioeconomic factors. For example, rural areas experienced a more notable increase in poverty compared to urban areas, possibly attributed to the generally lower economic resilience of rural households, rendering them susceptible to even minor economic hardships.[18] However, urban populations experienced greater employment loss and income reduction due to movement restrictions, overall, they suffered more economic shock than the rural.[18]

Thus, the few studies in Mozambique suggest that there had been an overall negative impact of the pandemic and associated lockdowns on the population. To further elucidate the prevalence of hardships experienced during the pandemic and examine potential risk factors for vulnerability to hardships, we used data collected by the Manhiça Health Research Center (CISM) through a cross-sectional household survey module added to the existing Health and Demographic Surveillance System (HDSS) in Manhiça district, Mozambique.

## METHODS

### Study design

This study is part of a broader examination within the Child Health and Mortality Prevention Surveillance (CHAMPS) Network to analyze the consequences of COVID-19 lockdowns for child health and mortality.[40-43] Leveraging the established platform within the CHAMPS Network of HDSS [44] in 2021, we administered a questionnaire to households in Manhiça, Mozambique.

### Study Setting

Manhiça is a district in Maputo Province, located about 85 km north of the country’s capital city, Maputo. An HDSS was established there in 1996 by CISM and currently covers the entire Manhiça District, which spans 2,380 km^2^ (Figure 1). More information about the Manhiça HDSS has been published elsewhere.[45, 46] With a population of 201,845, Manhiça is the second most populated district in Maputo Province after the Matola District (the capital of Maputo Province).[45] The population is mainly peasants and vendors, with a portion also working in two sugar factories in the district, namely Maragra, and Xinavane, which are the country’s primary sugar factories. The health system in Manhiça comprises one district hospital located in Manhiça village, one rural hospital located in the Xinavane administrative post, and 19 health centers.

**Figure 1.**
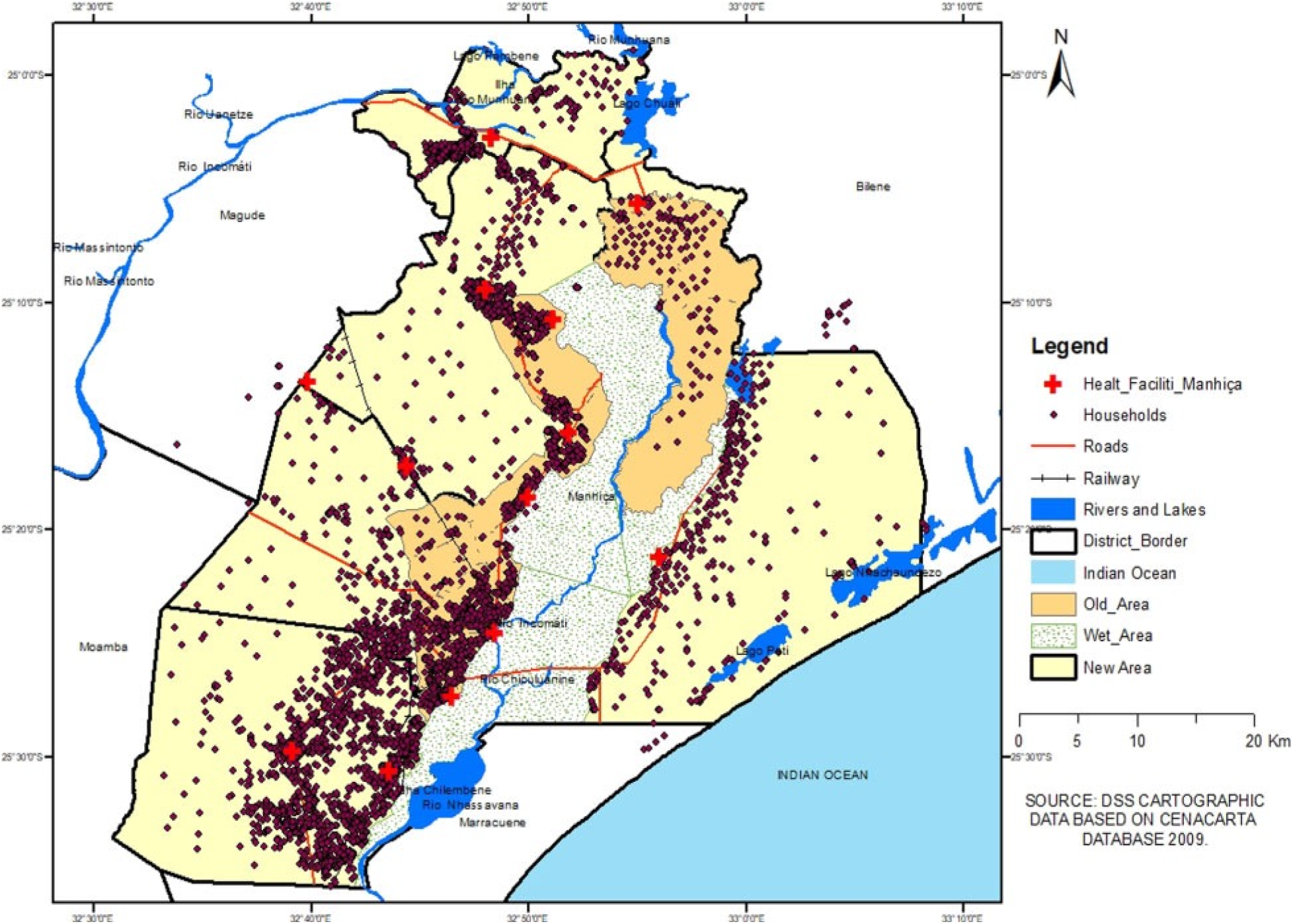
Map of Manhiça District. The original HDSS catchment area is shown in orange, a more recently expansion area is shown in yellow (Source: Created by the authors using HDSS cartographic data).

Manhiça district is a geographic corridor highly exposed to imported diseases such as HIV, and currently COVID-19, stemming from population migration. The district is intersected by Mozambique’s main road (National Road Number 1), which connects the Southern, Central, and Northern regions of the country as well as neighboring countries of South Africa, Eswatini, Zimbabwe, Malawi, Zambia, and Tanzania. The district is also crossed by a railway line connecting Zimbabwe to Maputo city and harbor, facilitating population mobility and disease spread. A large proportion of the Manhiça population migrate to the nearby capital city of Maputo or beyond, to South Africa and Eswatini, with regular return visits to their households in Manhiça.

### Study Participants

Study participants comprised heads of households or their representatives, defined as any household member aged 18 years or older who possessed adequate information about the household and its co-residing members, including visitors who had entered the household since March 2020 until the date of interview. All households within the Manhiça HDSS were eligible to participate in the study, aligning with the procedures and schedule of the HDSS rounds visits, during which this questionnaire was administered. This study adopted the definition of household and of household members used by the HDSS, defining a household as a group of one or more people living together in the same houses or group of houses within Manhiça District who share domestic expenses, eat together, and recognize one of them as their leader or representative. A member of the HDSS is a person who has resided in a household within Manhiça District for three or more months, or entered the study area with the intention of residence.[45]

### Survey Instrument

A survey instrument was developed to collect information about households’ experiences during the COVID-19 lockdown. It included information on knowledge regarding COVID-19 symptoms, transmission, and prevention; COVID-19 related shocks and household coping strategies; needs for and access to healthcare services for children under five years of age and for pregnant women; and food availability. The development of the survey followed state-of-the-art survey design methods. Steps in the design process included identifying central research concepts and questions; engaging in a rapid literature review; developing, adopting, and revising survey questions based on example questionnaires such as the one that the World Bank developed to assess economic and social consequences of the pandemic (i.e., the “High-Frequency Mobile Phone Surveys of Households to Assess the Impacts of COVID-19” survey); and pretesting the survey in the field.[47-49] Health authorities in the Manhiça district contributed to adapting the survey to local contexts and suggested additional questions to include in the instrument. The survey instrument prompted respondents to consider their household’s experiences since March 2020 while answering questions related to hardships that their household may have experienced during the pandemic. Data were collected electronically using tablets and a version of the survey that was translated into Portuguese.

Data collection was conducted by HDSS fieldworkers during their regular visits to the households through tablet-based in-person interviews with heads of households or their representatives if the heads were unavailable. Households were recruited in April 2021 before the Delta variant wave to the peak of Omicron cases in February 2022. In 2021, the HDSS recorded a total of 40,636 active households, of which 33,087 (81%) responded to the lockdown survey module. Data from the lockdown module were linked with data from the HDSS questionnaires, providing socio-demographic data about the household and their heads. Household variables included, among others: household size, number of children under 5 years of age, number of adults over 60 years, and number of pregnant women; household assets; and materials used for constructing the houses (Supplemental Materials, Figure 5). Variables about the head of household were age, sex, occupation, education, religion, ethnicity, and marital status.

The fieldworkers and their supervisors were trained by the study coordinator, a senior demographer at CISM, who led the adaptation and implementation of the Network module to Manhiça HDSS. Data cleaning and quality assurance followed standard procedures for the HDSS site, which covered every phase of the research process. In this process, 5% of households visited each week were revisited to confirm the recorded information.[45, 46, 50] Also, a script was used to filter errors at the time of uploading the data from tablets to the server, with records showing inconsistencies being returned to the field for reconciliations.

### Measures

The primary outcome variables of interest included five indicators of hardships a household reported experiencing since the onset of the COVID-19 pandemic (i.e., during the 1 to 1½ years since March 2020).^1^ Information on these household hardships were gathered from the following survey question: “Has your household been affected by any of these events since mid-March?” Responses included job loss, non-agricultural business closure, increases in the price of the main food products consumed, increases in the price of agricultural or commercial inputs, and having a household member detained.

Right-hand side variables. Head of household variables were age (coded as 1 = less than 30 years (reference), 2 = 30 to 39 years, 3 = 40 to 49 years, 4 = 50 to 59 years, 5 = 60 to 69 years, and 6 = 70+ years); sex (coded as male = 0 and female = 1); religion (coded as 1 = Zion (reference), 2 = Evangelical, 3 = Christian (non-denominational), 4 = Protestant, 5 = Catholic, 6 = other); ethnicity, defined by mother language,[51] (coded as 1 = Tsonga (reference), 2 = Sena/Ndau, 3 = Lomwe/Chuwabo 4 = Macua and 5 = European, 6 = other); education (coded as 1 = Primary (reference), 2 = Secondary, 3 = Technical, 4 = Tertiary, 5 = Literate, 6 = Other); and occupation (coded as 1 = farmer/fisherman/subsistence (reference), 2 = locksmith/bricklayer, 3 = guards/security, 4 = miner/factory worker, 5 = carpenter/electrician, 6 = lumberjack, 7 = merchants, 8 = health professional, 9 = military/policy, 10 = teacher, 11 = disabled/unemployed, 12 = volunteer, and 13= other).

Household variables included household size (coded as 1 = 1-2 individuals (reference), 2 = 3-4 individuals, 3 = 5-6 individuals, 4 = 7+ individuals); children under age 5 (coded as 0, 1+ individuals); adults over age 60 (coded as 0, 1+ individuals); and a wealth index based on a list of household assets (coded as 1 = poorest (reference), 2 = poorer, 3 = middle, 4 = richer, 5 = richest (see Appendix A for details of computation of this index). Specific household assets included vehicles (coded as 0 = none (reference), 1 = car, 2 = motorcycle, 3 = both); media (coded as 0 = none (reference), 1 = radio, 2 = tv, 3 = both).

### Analytic Strategy

Data cleaning and analysis was performed using R version 4.2.0.[37] Missing values for age, sex, religion, ethnicity, education, and occupation were less than 5 percent of the total observations. Descriptive statistics were reported for head of household and household characteristics. A series of un-adjusted logistic regression models were used to analyze associations between head of household and household characteristics with specific household hardships (i.e., business closure, food price increase, household member detained, input inflation, and job loss). Adjusted logistic regression models were then used to control for additional variables, which were included in the final adjusted model if their inclusion had theoretical justification. We assessed model fit with a comparison of Akaike Information Criterion (AIC) and Bayesian Information Criterion (BIC) scores—the smallest scores were for the fully adjusted model, indicating that this model had superior fit compared to simpler models. Results from the final logistic regression model are reported as Adjusted Odds Ratios (AOR) with 95% confidence intervals (95% CI) and visualized with forest plots.[52, 53]

Models were first run using the complete data set that included 33,087 households. A subsequent analysis was run with only those households who reported having at least one family member employed or operating a business during the HDSS round prior to the observation period. These selection criteria were used in order to only evaluate the 28,771 households that were at risk of having a family member lose a job, have a business close, or have prices increase in the inputs for their business or agricultural activities. Frequency distributions of the number of hardships experienced by a given household were comparable for the data with 33,087 households and 28,771 households. Likewise, results from the analyses using the complete data set and the analysis using the sub-sample were generally comparable. Hereafter, we present the results from the analysis of the sub-sample, results from the analysis using the complete data are available upon request.

## RESULTS

A majority of households (52.0%) were headed by a male household member and the majority (54.3%) of household heads were married (Table 1). The median and modal age category of the household head was 40-49 years. Tsonga was the most common ethnic group (92.3%); Sena/Ndau and Lomwe/Chuwabo combined represented about 3.1% of households. Roughly 91.4% of household heads identified religiously as either Zion, Evangelical, non-denominational Christian, Protestant, or Catholic. A majority (54.1%) of household heads had primary education, and 15.6% had secondary education. The most common occupations were farmer/fisherman/subsistence (53.1%) and miner/factory worker (13.1%); a little less than 6% were disabled or unemployed. Median household size was 3-4 household members. Of all households, 43.6% had children under 5 years and about 29.0% had adult family members over the age of 60. The percentage of households that reported a given hardship is presented in Figure 2; a Euler diagram presenting the shared distribution of hardships experienced by households during the pandemic is presented in Figure 3. A majority of households (59.2%) reported that they experienced 1 hardship during the observation period and another 18.2% reported experiencing more than 1 hardship. A little less than 25% of households reported that they did not experience any of the measured hardships during the observation period (Figure 2).

**Table 1:** Descriptive Statistics for Head of Household Characteristics (n=28,771)

|  |  | n | % |
| --- | --- | --- | --- |
| **Head of Household Characteristics** |  |  |  |
| Age | <30 | 2513 | 8.70% |
|  | 30-39 | 6561 | 22.71% |
|  | 40-49 | 7103 | 24.59% |
|  | 50-59 | 4843 | 16.76% |
|  | 60-69 | 3797 | 13.14% |
|  | 70+ | 3643 | 12.61% |
| Sex | Female | 15013 | 51.96% |
|  | Male | 13608 | 47.10% |
| Marital Status | Single | 2684 | 9.29% |
|  | Married | 15694 | 54.32% |
|  | Divorced/Separated | 3506 | 12.14% |
|  | Widowed | 6202 | 21.47% |
| Religion | Zion | 9613 | 33.27% |
|  | Evangelical | 3868 | 13.39% |
|  | Christian (Non-denominational) | 3276 | 11.34% |
|  | Protestant | 6147 | 21.28% |
|  | Catholic | 2629 | 9.10% |
|  | Other | 2477 | 8.57% |
| Ethnicity | Tsonga | 26658 | 92.27% |
|  | Sena/Ndau | 441 | 1.53% |
|  | Lomwe/Chuwabo | 440 | 1.52% |
|  | Macua | 129 | 0.45% |
|  | European | 95 | 0.33% |
|  | Other | 284 | 0.98% |
| Education | Primary | 15620 | 54.07% |
|  | Secondary | 4511 | 15.61% |
|  | Technical | 239 | 0.83% |
|  | Tertiary | 299 | 1.03% |
|  | Literate | 954 | 3.30% |
|  | Other | 2810 | 9.73% |
|  | No Education | 3325 | 11.51% |
| Occupation | Farmer/Fisherman/Subsistence | 15330 | 53.06% |
|  | Locksmith/Bricklayer | 994 | 3.44% |
|  | Guards/Security | 530 | 1.83% |
|  | Miner/Factory Worker | 3787 | 13.11% |
|  | Carpenter/Electrician | 128 | 0.44% |
|  | Lumberjack | 222 | 0.77% |
|  | Merchants | 1770 | 6.13% |
|  | Health Professional | 198 | 0.69% |
|  | Military/Policy | 261 | 0.90% |
|  | Teacher | 425 | 1.47% |
|  | Disabled/Unemployed | 1673 | 5.79% |
|  | Volunteer | 420 | 1.45% |
|  | Other | 2322 | 8.04% |

Notes: Data source: CHAMPS Mozambique Lockdown Module data and CHAMPS Mozambique Health and Demographic Surveillance data. Less than 5 percent of observations were missing for any of the variables for head of household characteristics. The original data set, representing 33,087 households, was restricted to only those households that had reported at least one family member employed or engaged in business or agricultural activities in the HDSS round prior to the observation period; this resulted in a subset of data that included 28,771 households.

**Table 1.1:** Descriptive Statistics for Household Characteristics (n=28,771)

|  |  | n | % |
| --- | --- | --- | --- |
| **Household Characteristics** |  |  |  |
| Household Size | 1-2 | 6919 | 23.95% |
|  | 3-4 | 8403 | 29.09% |
|  | 5-6 | 7802 | 27.00% |
|  | 7+ | 5647 | 19.55% |
| Children Under Age 5 | 0 | 16163 | 55.94% |
|  | 1+ | 12608 | 43.64% |
| Adults Over Age 60 | 0 | 20385 | 70.56% |
|  | 1+ | 8386 | 29.03% |
| Wealth Index | Richest | 5888 | 20.38% |
|  | Richer | 5749 | 19.90% |
|  | Middle | 5897 | 20.41% |
|  | Poorer | 5635 | 19.50% |
|  | Poorest | 5602 | 19.39% |
| Media | None | 12733 | 44.07% |
|  | Computer | 3401 | 11.77% |
|  | TV | 6551 | 22.67% |
|  | Both | 6086 | 21.07% |
| Vehicles | None | 26523 | 91.80% |
|  | Car | 1553 | 5.38% |
|  | Motorcycle | 510 | 1.77% |
|  | Both | 185 | 0.64% |

Notes: Data source: CHAMPS Mozambique Lockdown Module data and CHAMPS Mozambique Health and Demographic Surveillance data. No observations were missing for any of the variables for household characteristics. The original data set, representing 33,087 households, was restricted to only those households that had reported at least one family member employed or engaged in business or agricultural activities in the HDSS round prior to the observation period; this resulted in a subset of data that included 28,771 households.

**Figure 2.**
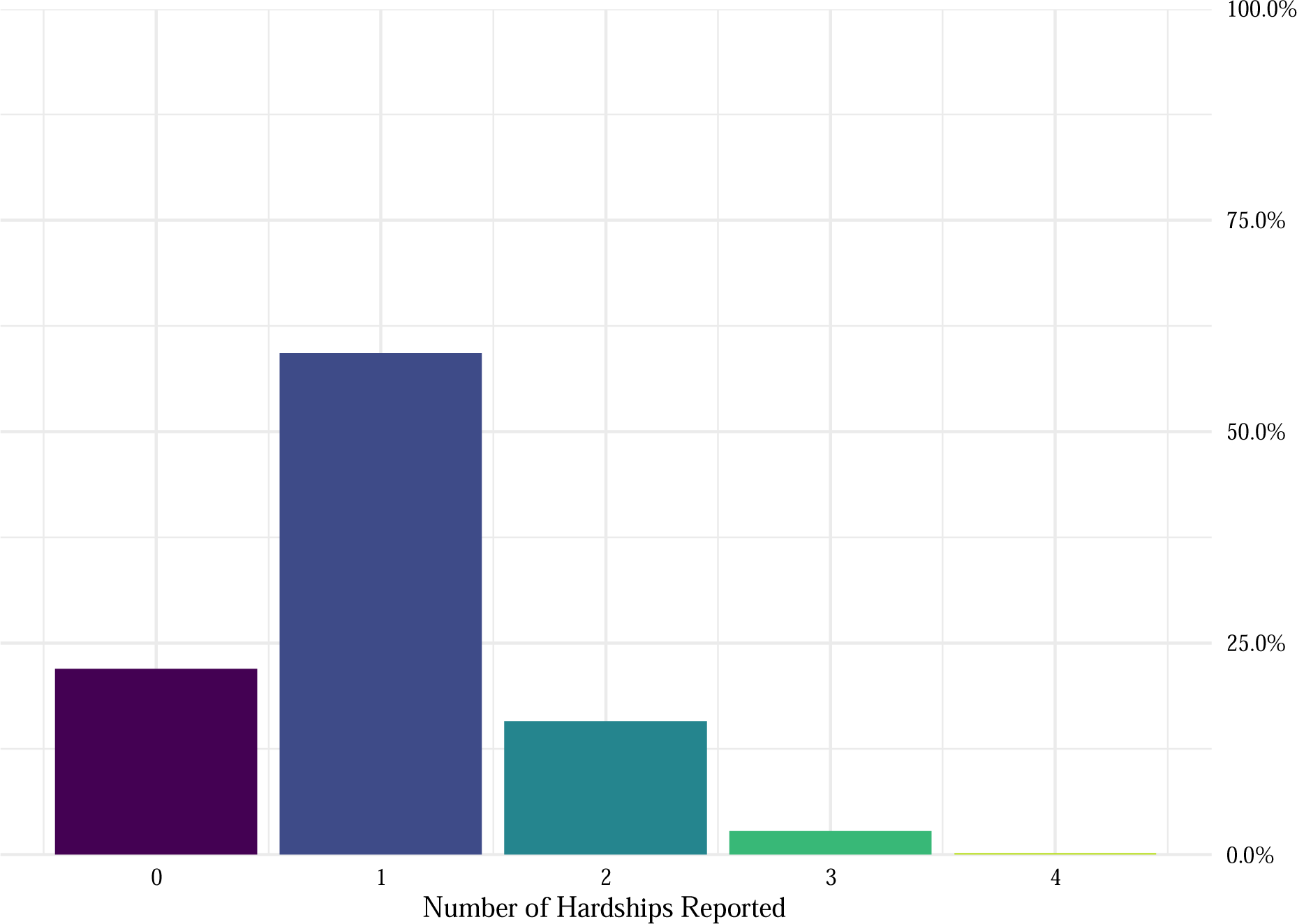
Percentage of Households that Reported a Given Number of Hardships (n = 28,711)

**Figure 3.**
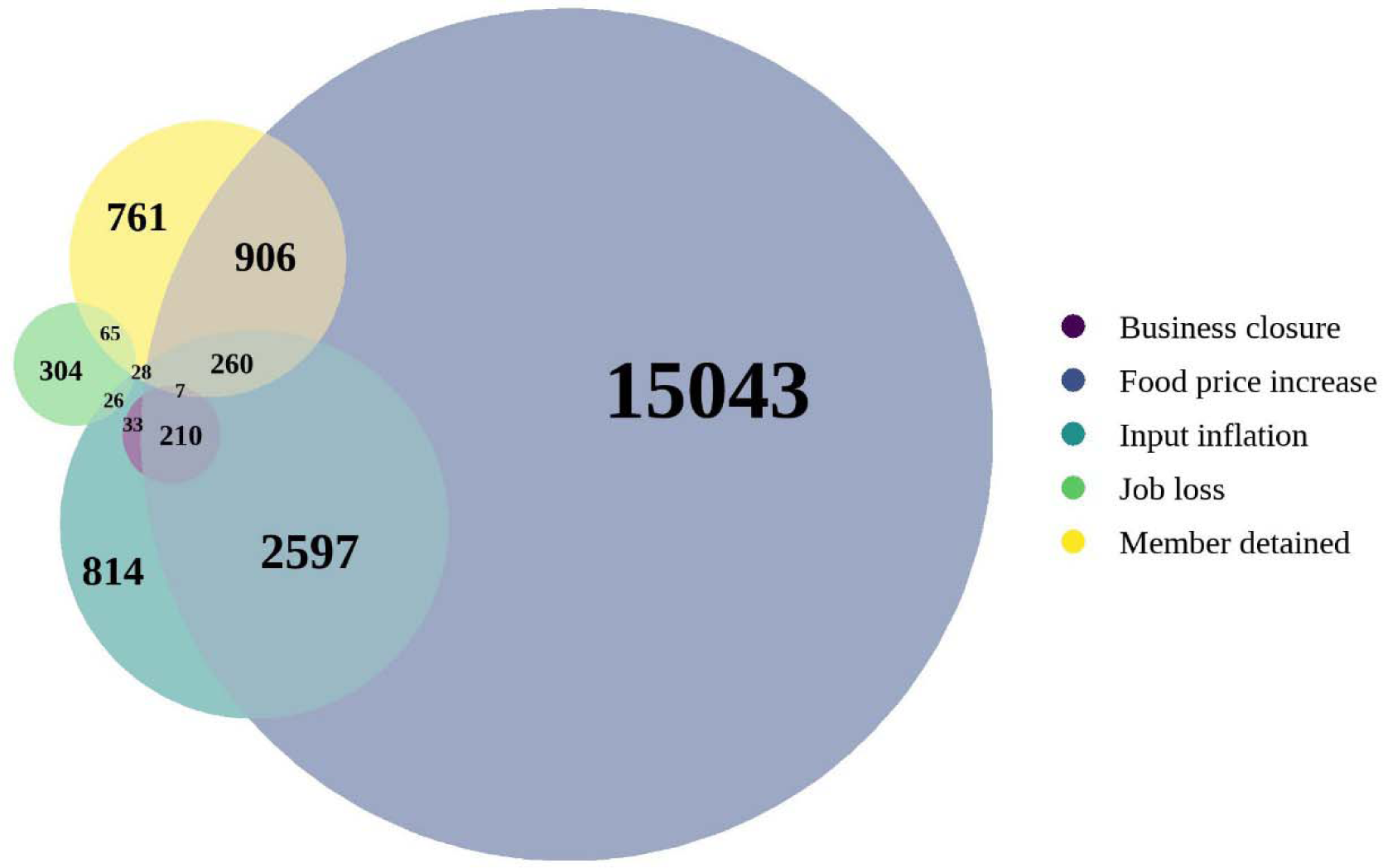
Pattern of Multiple Hardships Reported by Households (n = 28,711). Euler diagram representing the overlap of hardships that households reported experiencing during the pandemic.

### Price Increases for Food

The most common hardship that households reported experiencing during the observation period was increases in food prices, with 70.3% of households reporting this hardship (Figure 4). After adjusting for other variables (see Figure 5 for a composite summary of results for all 5 hardships; detailed results are provided in supplementary figures in Appendix B), households were more likely to report increases in food prices if their household head identified as Protestant (AOR = 1.73, 95% CI [1.60, 1.77]) or Catholic (AOR = 1.27, 95% CI [1.15, 1.41]) compared to Zion or if the head of household was divorced or separated compared to single (AOR = 1.27, 95% CI [1.15, 1.41]). Compared to households whose heads worked in farming, fishing, or subsistence activities, households were more likely to report increases in food prices if their household head was employed as a locksmith or bricklayer (AOR = 1.29, 95% CI [1.05, 1.59]), a carpenter or electrician (AOR = 1.35, 95% CI [1.01, 1.83]), a merchant (AOR = 1.27, 95% CI [1.12, 1.44]), in the military or police force (AOR = 1.88, 95% CI [1.36, 2.66]), as a teacher (AOR = 1.43, 95% CI [1.09, 1.87]), or if they were unemployed or disabled (AOR = 1.79, 95% CI [1.57, 2.04]). Households with a larger number of household members were more likely to report increases in food prices; for example, compared to households with 1-2 members, the odds that a household reported increases in food prices were higher if the household had 3-4 members (AOR = 1.20, 95% CI [1.11, 1.29]), 5-6 members (AOR = 1.24, 95% CI [1.13, 1.35]), or 7 or more members (AOR = 1.21, 95% CI [1.10, 1.34]). Households were also more likely to report increases in food prices if they owned a TV (AOR = 1.79, 95% CI [1.65, 1.95]) or both TV and radio (AOR = 1.76, 95% CI [1.61, 1.91]).

**Figure 4.**
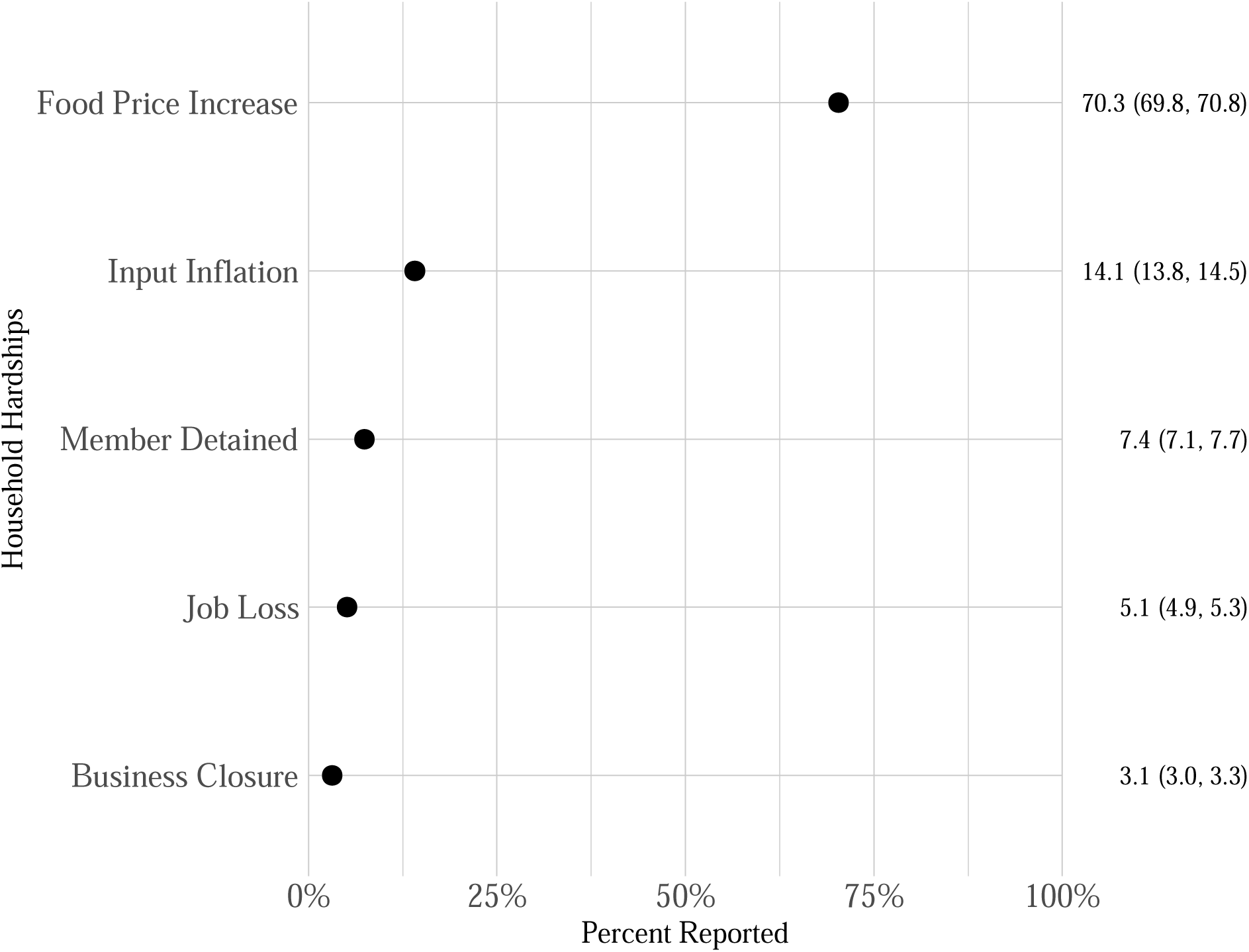
Percentage (and 95% confidence interval) of Households that Reported a Given Hardship (n = 28,711).

**Figure 5.**
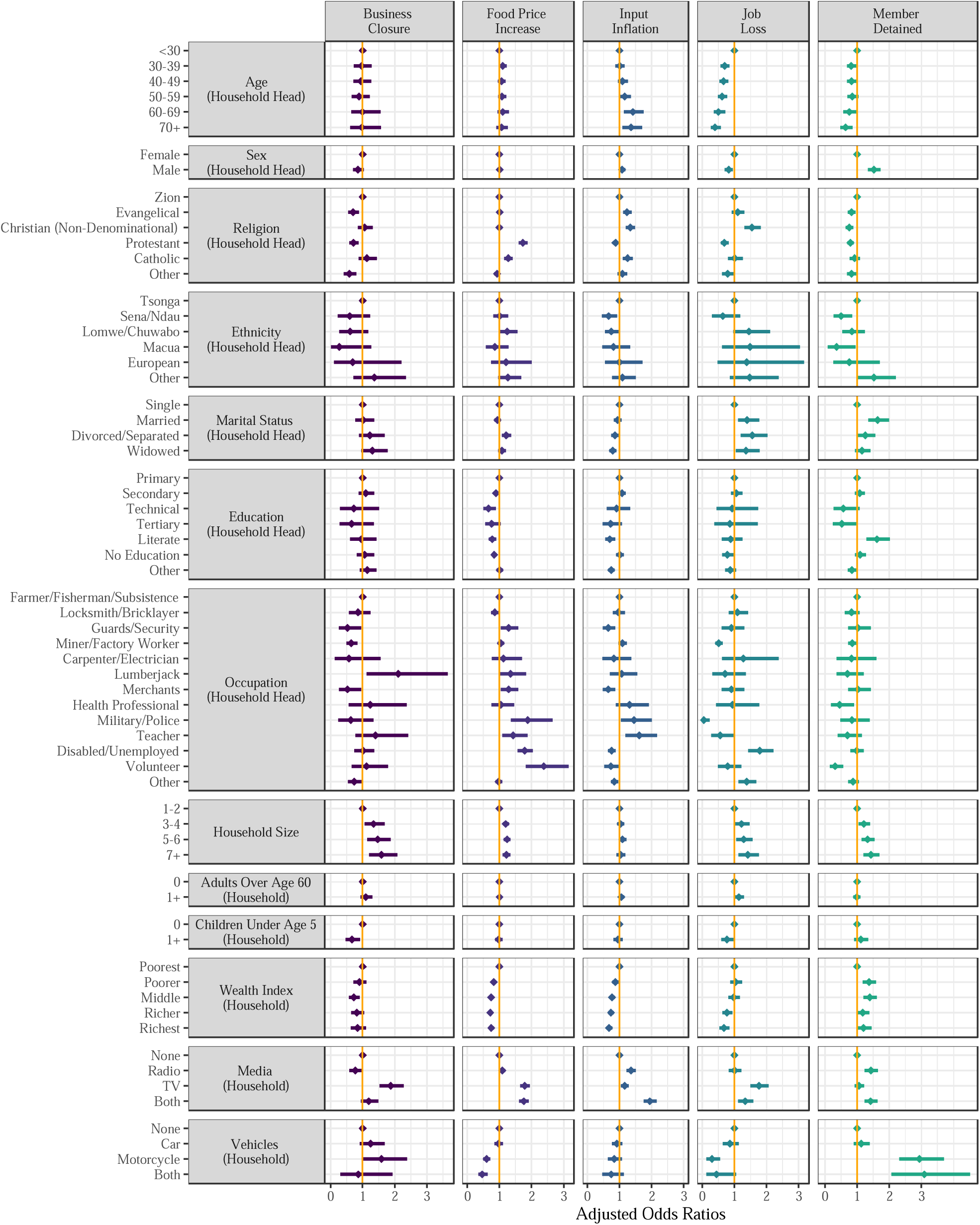
Adjusted Associations between Individual and Household Characteristics with Hardships Reported by Households Living in Manhiça, Mozambique (n=27,404). Presented as Adjusted Incidence Risk Ratios with 95% confidence intervals. Missing values for age, sex, religion, ethnicity, education, and occupation were less than 5 percent of the total observations.

Households were generally less likely to report increases in food prices if their household head had a higher level of education, were relatively wealthier, or if they had access to means of transportation. After adjusting for other variables, households were less likely to report increases in food prices during the pandemic if the head of household had a secondary education (AOR = 0.89, 95% CI [0.82, 0.97]), a technical education (AOR = 0.66, 95% CI [0.50, 0.90]), a technical education (AOR = 0.66, 95% CI [0.50, 0.90]), a tertiary education (AOR = 0.76, 95% CI [0.56, 1.04]), or identified as being literate (AOR = 0.78, 95% CI [0.67, 0.90]) compared having only a primary education, although the association with having a tertiary education was not statistically significant. Compared to the poorest households, households were less likely to report food price increases during the pandemic, after adjusting for other variables, if the household fell within the poorer (AOR = 0.82, 95% CI [0.76, 0.90]) to richest (AOR = 0.75, 95% CI [0.67, 0.83]) wealth index quintiles. Households were less likely to report food prices increases if they owned a motorcycle (AOR = 0.60, 95% CI [0.50, 0.72]) or both a motorcycle and a car (AOR = 0.47, 95% CI [0.34, 0.64]).

### Price Increases for Agricultural or Business Inputs

After increases in food prices, the next most common hardship that households reported experiencing was increases in the prices of agricultural or business inputs (14.1% of households reported this hardship). After adjusting for other variables (see Figure 5), age of the household head was positively associated with increases in the price of agricultural or business inputs—compared to a household head younger than 30 years, households were more likely to have experienced input increases if they were headed by someone who was age 60-69 (AOR = 1.41, 95% CI [1.14, 1.75]) or 70 years old or more (AOR = 1.36, 95% CI [1.09, 1.71]). Households were also more likely to report increases in the price of agricultural or business inputs if their household head identified as Evangelical (AOR = 1.73, 95% CI [1.60, 1.77]), non-denominational Christian (AOR = 1.73, 95% CI [1.60, 1.77]) or Catholic (AOR = 1.27, 95% CI [1.15, 1.41]) compared to the Zion denomination. Increases in the price of agricultural or business inputs were likewise associated with household ownership of a radio (AOR = 1.36, 95% CI [1.21, 1.52]), a TV (AOR = 1.16, 95% CI [1.04, 1.29]) or both TV and radio (AOR = 1.95, 95% CI [1.75, 2.16]). In contrast, household wealth status was negatively associated with increases in the price of agricultural or business inputs; compared to the poorest households, households were less likely to report increases in the price of agricultural or business inputs, if the household fell within the poorer (AOR = 0.88, 95% CI [0.79, 0.97]) to richest (AOR = 0.68, 95% CI [0.59, 0.78]) wealth index quintiles.

### Household Member Detained

About 7.4 percent of households reported having a household member detained (e.g., prevented from returning home due to mobility restraints) during the observational period. Households were more likely to report experiencing this if they were headed by a female (AOR = 1.52, 95% CI [1.34, 1.73]); had a household head who was married; had households with 7 or more members (AOR = 1.43, 95% CI [1.20, 1.45])) compared to 1-2 members; were from the poorer (AOR = 1.37, 95% CI [1.18, 1.60]) or middle (AOR = 1.39, 95% CI [1.20, 1.62]) wealth quintiles compared to the poorest; owned a radio (AOR = 1.43, 95% CI [1.23, 1.65]) or both TV and radio (AOR = 1.42, 95% CI [1.23, 1.63]); or if they owned a motorcycle (AOR = 2.94, 95% CI [2.31, 3.71]) or both a motorcycle and a car (AOR = 3.10, 95% CI [2.07, 4.52]). Households were less likely to have experienced having a family member detained if they were headed by someone who was 70 years old or more (AOR = 0.64, 95% CI [0.48, 0.87]) compared to a household head younger than 30 years. Households were less likely to have experienced having a family member detained if their household head identified as Evangelical (AOR = 0.83, 95% CI [0.71, 0.96]), non-denominational Christian (AOR = 0.75, 95% CI [0.64, 0.88]) or Catholic (AOR = 0.79, 95% CI [0.69, 0.90]) compared to the Zion denomination.

### Household Member Job Loss

A little over five percent of households reported having a household member lose their job during the pandemic period. Households were at greater risk of having a family member lose their job if the household head was non-denominational Christian (AOR = 1.55, 95% CI [1.31, 1.83]) compared to Zion; married (AOR = 1.40, 95% CI [1.11, 1.75]), divorced or separated (AOR = 1.56, 95% CI [1.20, 2.04]), or widowed (AOR = 1.37, 95% CI [1.05, 1.79]) compared to single; had larger households (e.g., 7 or more members (AOR = 1.42, 95% CI [1.13, 1.77])) compared to 1-2 members); or owned a TV (AOR = 1.76, 95% CI [1.50, 2.07]) or both TV and radio (AOR = 1.33, 95% CI [1.12, 1.59]). Conversely, the age of the household head was negatively associated with job loss; e.g., compared to a household head younger than 30 years, households were less likely to have experienced having a family member lose their job if they were headed by someone who was age 30-39 (AOR = 0.69, 95% CI [0.57, 0.84]), 40-49 (AOR = 0.66, 95% CI [0.54, 0.81]), 50-59 (AOR = 0.62, 95% CI [0.49, 0.78]), 60-69 (AOR = 0.51, 95% CI [0.36, 0.72]), or 70 years old or more (AOR = 0.40, 95% CI [0.27, 0.58]). Head of household employment in the military or police force, compared to farming, fishing, or subsistence, was negatively associated with having a household member lose their job (AOR = 0.05, 95% CI [0.00, 0.24]). Households were also less likely to report job loss if the household was in the richer (AOR = 0.77, 95% CI [0.63, 0.94]) or richest (AOR = 0.68, 95% CI [0.54, 0.85]) wealth index quintiles. Likewise, households were less likely to report having a household member lose their job if they owned a motorcycle (AOR = 0.30, 95% CI [0.13, 0.56]) or both a motorcycle and a car (AOR = 0.44, 95% CI [0.13, 1.05]); however, the association between ownership of both a motorcycle and a car and job loss was not statistically significant.

### Business Closure

The least commonly reported hardship by households was having a business closure (3.1%). Having a household head employed as a carpenter/electrician compared to farming, fishing, or subsistence was positively associated with business closure (AOR = 2.11, 95% CI [1.12, 3.65]). Larger households with 7 or more members were also more likely to have had a business closure (AOR = 1.58, 95% CI [1.20, 2.08]) compared to 1-2 members. Conversely, households were less likely to experience a business closure if the household head was employed as a security guard (AOR = 0.53, 95% CI [0.26, 0.96]) or miner/factory worker closure (AOR = 0.65, 95% CI [0.50, 0.84]) compared to farming, fishing, or subsistence. Having an adult household member over the age of 60 was likewise negatively associated with business closure (AOR = 0.67, 95% CI [0.48, 0.92]).

## DISCUSSION

In this study, we examined the prevalence of several household hardships during the COVID-19 pandemic in the district of Manhiça of Mozambique. We further examined the demographic, economic, and social characteristics of individuals and households associated with these hardships. Roughly 78.1% of households reported experiencing at least one hardship since the onset of the pandemic, the most commonly reported hardship was an increase in local food prices. In preliminary analyses, we considered evaluating additive and PCA based indices of these household hardships; however, we concluded that the combination of these variables resulted in a poor fit for an index after evaluating the additive index via Cronbach’s Alpha scores and the results of the PCA analysis. The results from our analyses further suggest that there was utility in evaluating each hardship independently as associations with individual and household level factors were heterogeneous across the hardships examined, particularly with regards to the factors associated with having a household member detained.

Individual-level risk factors for experiencing these hardships included head of household characteristics such as lower levels of education, a variety of occupations (merchants, health professionals, military/police, teachers, volunteers) as well as being disabled or unemployed. Household-level risk factors included having a larger household and having lower wealth status as indicated by ownership of fewer household assets. In general, the pattern of association for household wealth is suggestive of a dose-response; households with greater wealth (more assets) were at significantly lower risk for experiencing hardships. These associations are largely consistent with risk factors reported in studies from other resource-limited countries.[6,10-13,17] Overall, TV and radio ownership were positively associated with experiencing hardships. This positive association persisted even after to controlling for household wealth, suggesting that this relationship may be attributed to the functionality of these assets as a means of mass communication and not just as an indicator of wealth; i.e., households with a TV or radio may be better informed of the events surrounding the pandemic and fear what is happening in Maputo, South Africa and around the world. Household ownership of a motorcycle or both a motorcycle and a car were inversely related to experiencing hardships. This may relate to transportations’ ability to empower individuals and households to transcend local conditions and access resources from a larger geographic area, thereby increasing their resilience to hardships during the pandemic.[30-32] In contrast to prior findings,[18] having a head of household employed in farming, fishing, or subsistence was protective in some instances against households experiencing hardships compared to several other occupations. This observation is consistent with the local economic context of Manhiça, where farming dominates the lower Inkomati River region, which occupies large extensions of the district from north to the south. The agriculture in Manhiça generally consists of family-based subsistence farming, supplemented by some commercial farming, along with employment in sugar-cane plantations that are central to the region’s economy. Farming, fishing, or subsistence activities were not subject to lockdown measures, potentially contributing to the reduced impact of hardships on households engaged in these activities. Future interventions that seek to mitigate negative consequences associated with pandemic mitigation efforts should anticipate these characteristics as important risk factors and target interventions to aid vulnerable groups.

The empirical findings from this study are specific to the community living within the study site; a limitation common to studies using HDSS data.[16] However, our findings are generally consistent with findings reported in other studies from sub-Saharan African countries.[40-42] As an observational study, other potential limitations include unmeasured variable bias (for example, we did not gather information related to food insecurity beyond increases in local food prices) or recall bias due to the extended length of time considered in the study (i.e., some respondents may have forgotten hardships that occurred closer to the onset of the pandemic). We also cannot draw causal inferences given the cross-sectional nature of the study. Results from other studies suggest hardships associated with the COVID-19 pandemic are ephemeral.[7] Looking forward, scholars should collect and analyze longitudinal data to evaluate hardship events over time and explore potential causal relationships. Demographic surveillance systems present an efficient platform for follow-up data collection using the same survey instrument attached to subsequent rounds of regular data collection.

## CONCLUSION

Households in Manhiça experienced a variety of hardships during the COVID-19 pandemic, with the most prevalent hardship being the surge in local food prices. Restrictions of travel and closure of borders, particularly the border with South Africa, may have been the most important causes of this increase in food prices. Southern Mozambique relies heavily on imported food supplies from South Africa and certain provinces in Central and Northern Mozambique. Informal vendors of different scales play an important (if not the major) role in the amount of imported goods from South Africa. The price hikes commenced following the South African government’s announcement of border closures in March 2020, and even during periods of border reopening, travel to South Africa incurred additional expenses due to mandatory COVID-19 testing at the border. Households headed by individuals with lower education and employed in specific occupations as well as households that were larger in size or poorer in asset ownership compared to other households were generally at greater risk of experiencing a larger variety of hardships. Conversely, households that owned “distance demolishing technologies” such as motorcycles were less likely to experience several of these hardships, presumably because they were able to transcend local conditions.[29-32] Associations between specific hardships and individual and household characteristics varied to some degree; these variations highlight the importance of considering specific hardships when planning and implementing interventions to improve the health and well-being of populations living in these conditions. These findings may inform future policies and practices aiming to mitigate the negative consequences of COVID-19 and future disease outbreaks.

## DECLARATIONS

### Ethics approval and consent to participate

This study was conducted according to the guidelines in the Declaration of Helsinki; all procedures involving research study participants, including digital data collection using tablets that were programmed with the corresponding survey instruments, were approved by the Institutional Ethics Review Board for Health (CIBS) approval reference number No Ref:CIBS-CISM/15/2021. Written informed consent was obtained for participants who were able to read and write. For participants who were unable to read or write, the informed consent statement was read, and oral informed consent from the participant was obtained, documented, and witnessed. These procedures for obtaining written or oral informed consent were approved by the Institutional Ethics Review Board for Health (CIBS), a review board affiliated with the National Bioethics Committee for Health (CNBS).

### Consent for publication

All authors provide consent for publication.

### Availability of data and materials

The COVID-19 lockdown dataset analyzed for this study can be found in the CHAMPS Population Surveillance Dataverse [link].[54] The Manhiça HDSS data that was used in combination with the COVID-19 lockdown dataset are available from the authors upon reasonable request.

### Author contributions

Conceptualization (AN, CS, TM, QB, IM), data curation (AN, CS, TM, QB, IM), formal analysis (JM, ZM), investigation (AN, CS, TM, QB, IM), methodology (AN, ZM, JM, CS, TM, QB, IM, SC), project administration (AN, CS, TM, QB, IM, SC), resources (AN, CS, TM, QB, IM), supervision (AN, QB, IM, SC), visualization (ZM, JM), writing – original draft preparation (JM, ZM), writing – review & editing (AN, ZM, JM, CS, TM, QB, IM, SC).

## Acknowledgements

We are indebted to the households living within the Manhica HDSS that participated in our study and voluntarily contributed their time and energy in responding to our survey questionnaire in addition to ongoing participation in the regular data collection activities of the HDSS. We are also indebted to the HDSS team members who fielded the survey.

## SUPPLEMENTARY MATERIAL

## Appendix A. Household Wealth Index Generation

A household wealth index was generated using principal components analysis and a list of household assets and construction materials for the main dwelling of a given household. To generate this index, we followed the recommendations from the Demographic and Health Surveys Program (DHS) and the World Food Programme (WFP) that outline steps for calculating an asset-based wealth index,[55-57] including coding instructions for implementation in Stata,[58] and an adaptation of this coding process that we implemented in R in a prior study.[30, 32] Using these documents to guide us, we generated a wealth index for Manhiça households by identifying a list of household assets (see Figure A1). Assets were recoded into dichotomous variables representing whether or not a household owned a given asset; dwelling materials were recoded into improved vs. non-improved. The WFP recommends that a given variable should be included in subsequent calculations of the index only if the percent of ownership ranged between 5 and 95 percent. We employed principal components analysis with varimax rotation to calculate component scores for households, which explained 45% of the variation; extracted the PCA scores of the first component; and finally organized the scores into wealth quintiles to generate an asset index. The distribution of assets owned by households as well as the materials used for constructing a household’s residence are presented as percentages in Figure (A1).

**Figure A1.**
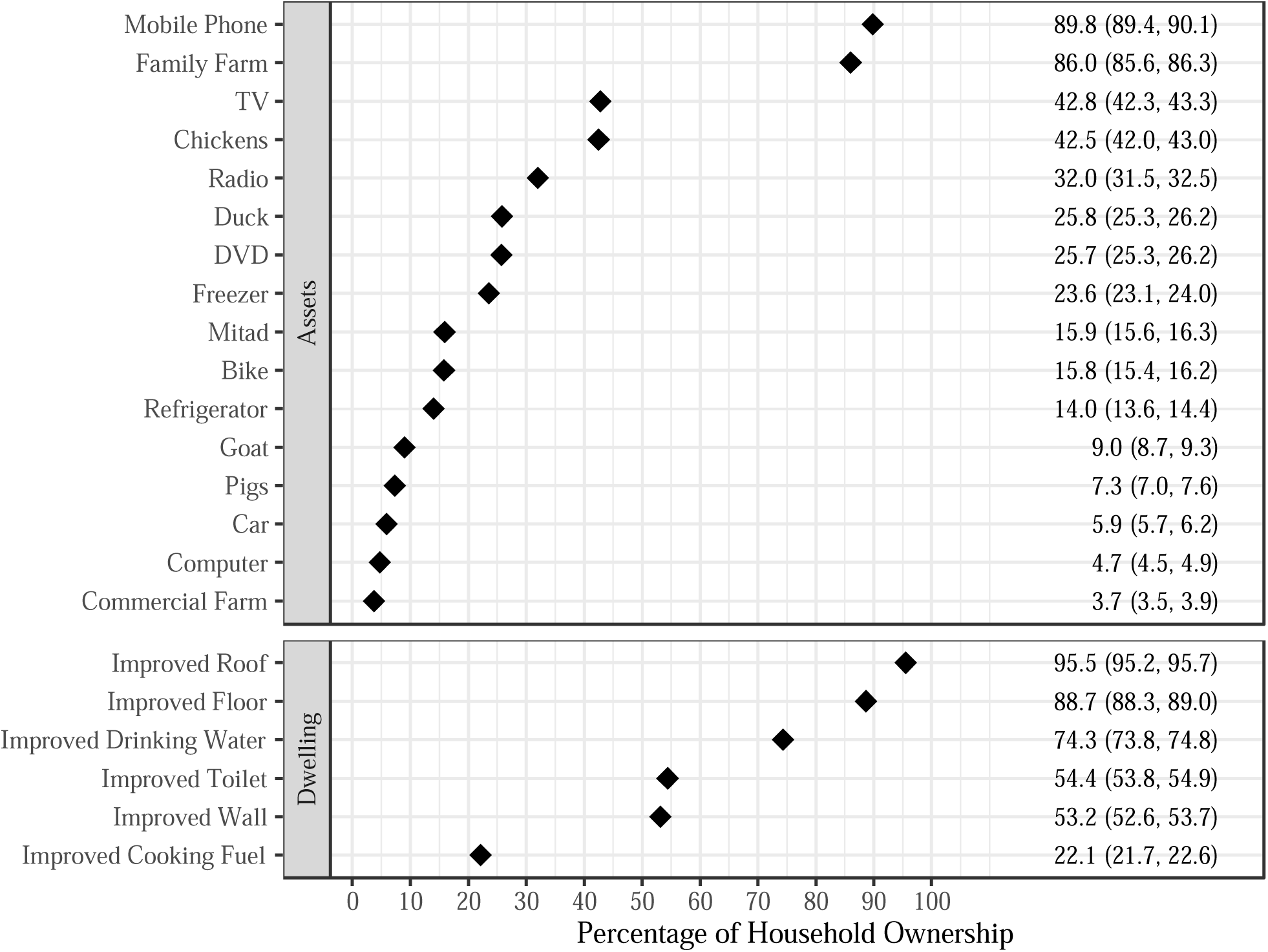
Household Asset Ownership and Dwelling Construction Materials. A Mitad is a portable, electric hotplate used for cooking.

## Appendix B. Hardship Specific Analyses

**Figure B.1.**
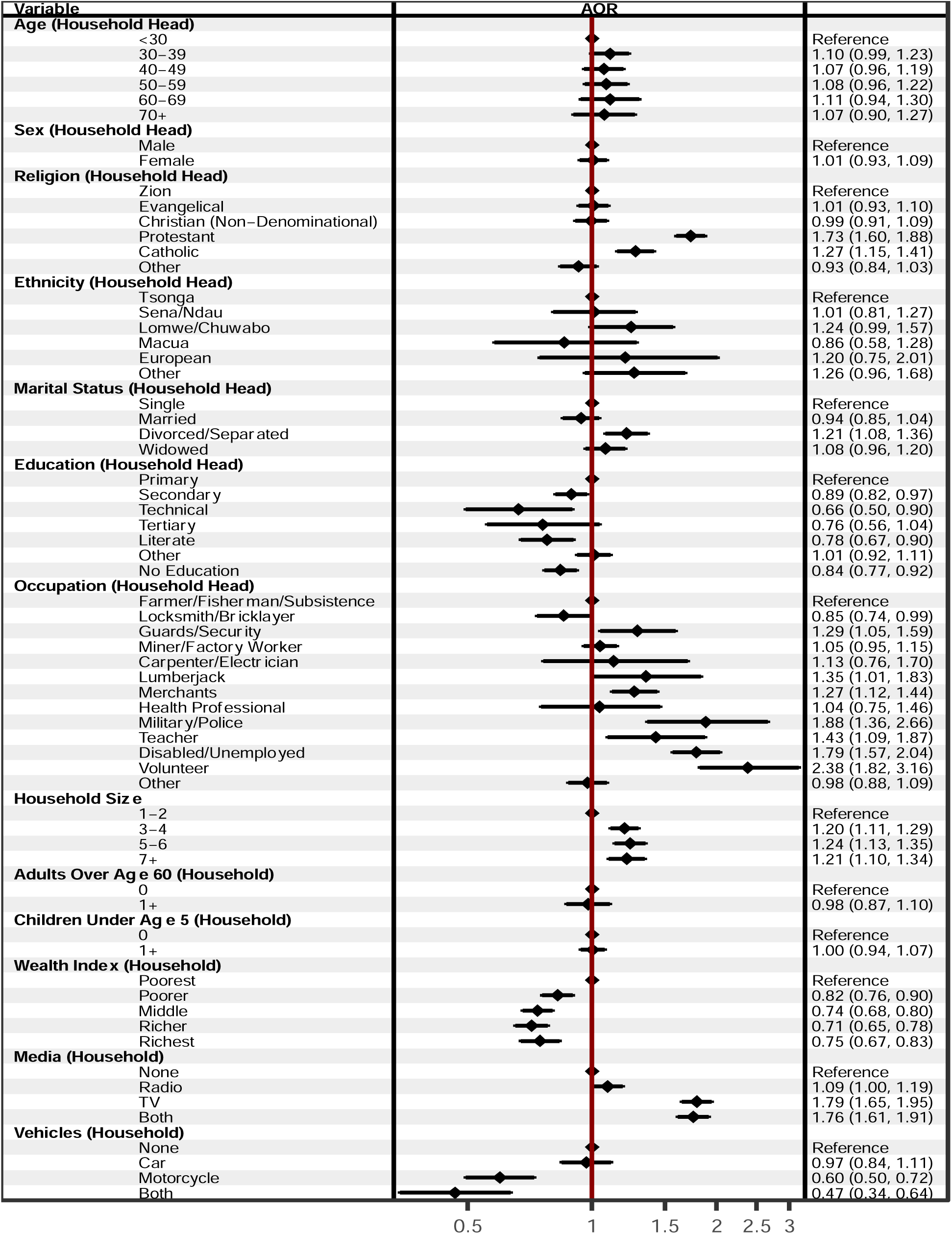
Adjusted Associations with Food Price Increases among Households Living in Manhiça (n=27,404). Presented as Adjusted Odds Ratios with 95% confidence intervals. Missing values for age, sex, religion, ethnicity, education, and occupation were less than 5 percent of the total observations.

**Figure B.2.**
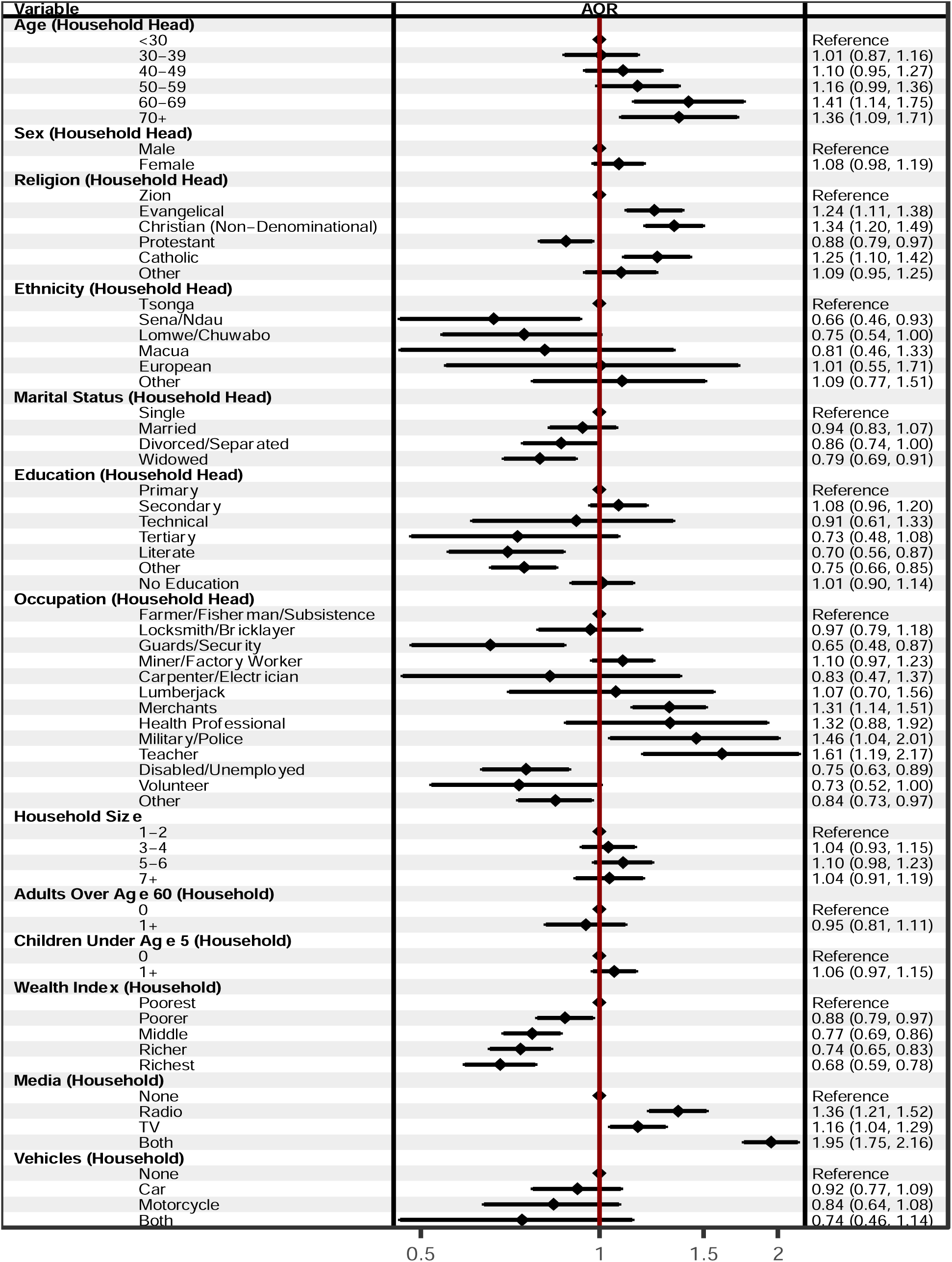
Adjusted Associations with Agricultural or Business Inputs Inflation among Households Living in Manhiça (n=27,404). Presented as Adjusted Odds Ratios with 95% confidence intervals. Missing values for age, sex, religion, ethnicity, education, and occupation were less than 5 percent of the total observations.

**Figure B.3.**
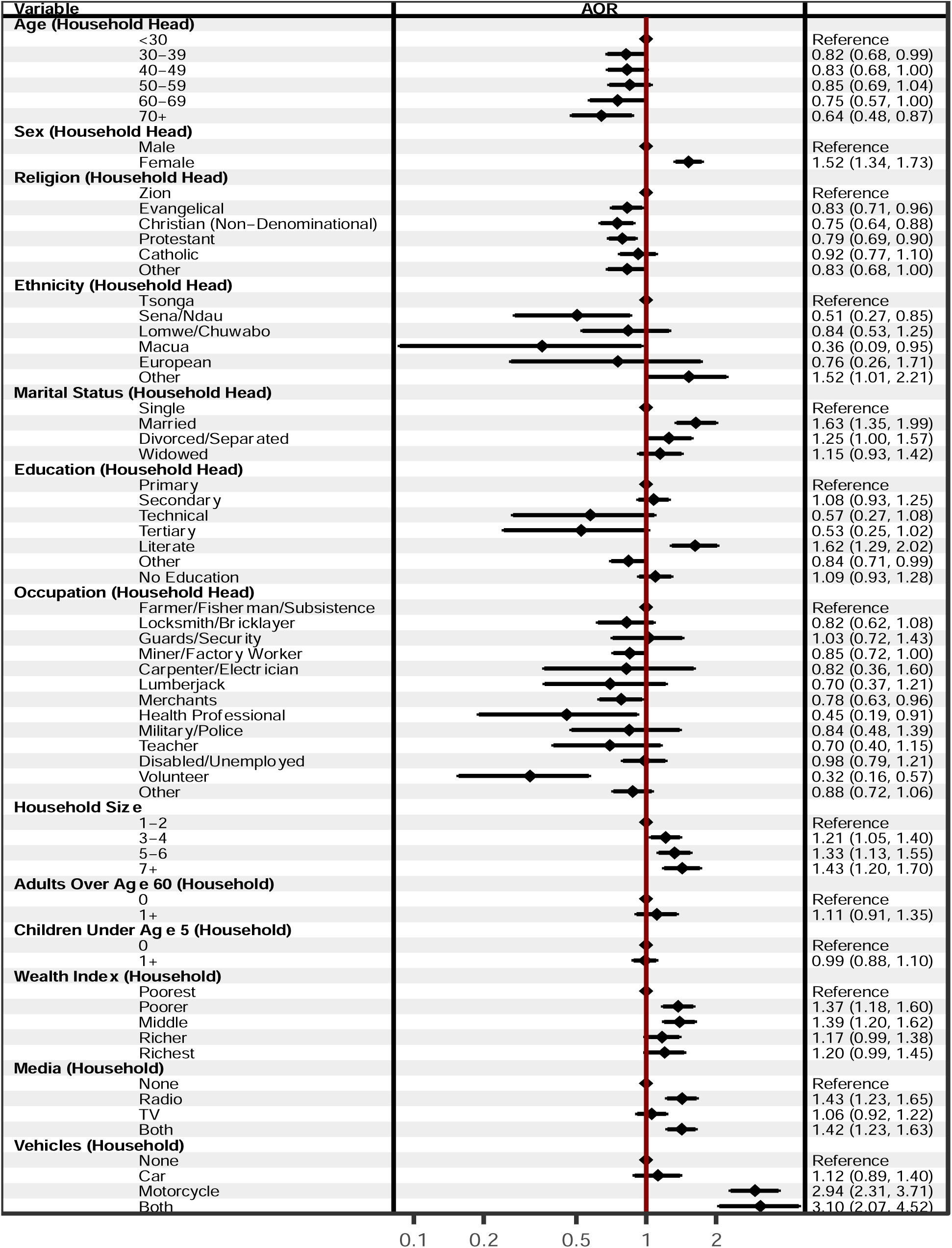
Adjusted Associations with having a Household Member Detained among Households Living in Manhiça (n=27,404). Presented as Adjusted Odds Ratios with 95% confidence intervals. Missing values for age, sex, religion, ethnicity, education, and occupation were less than 5 percent of the total observations.

**Figure B.4.**
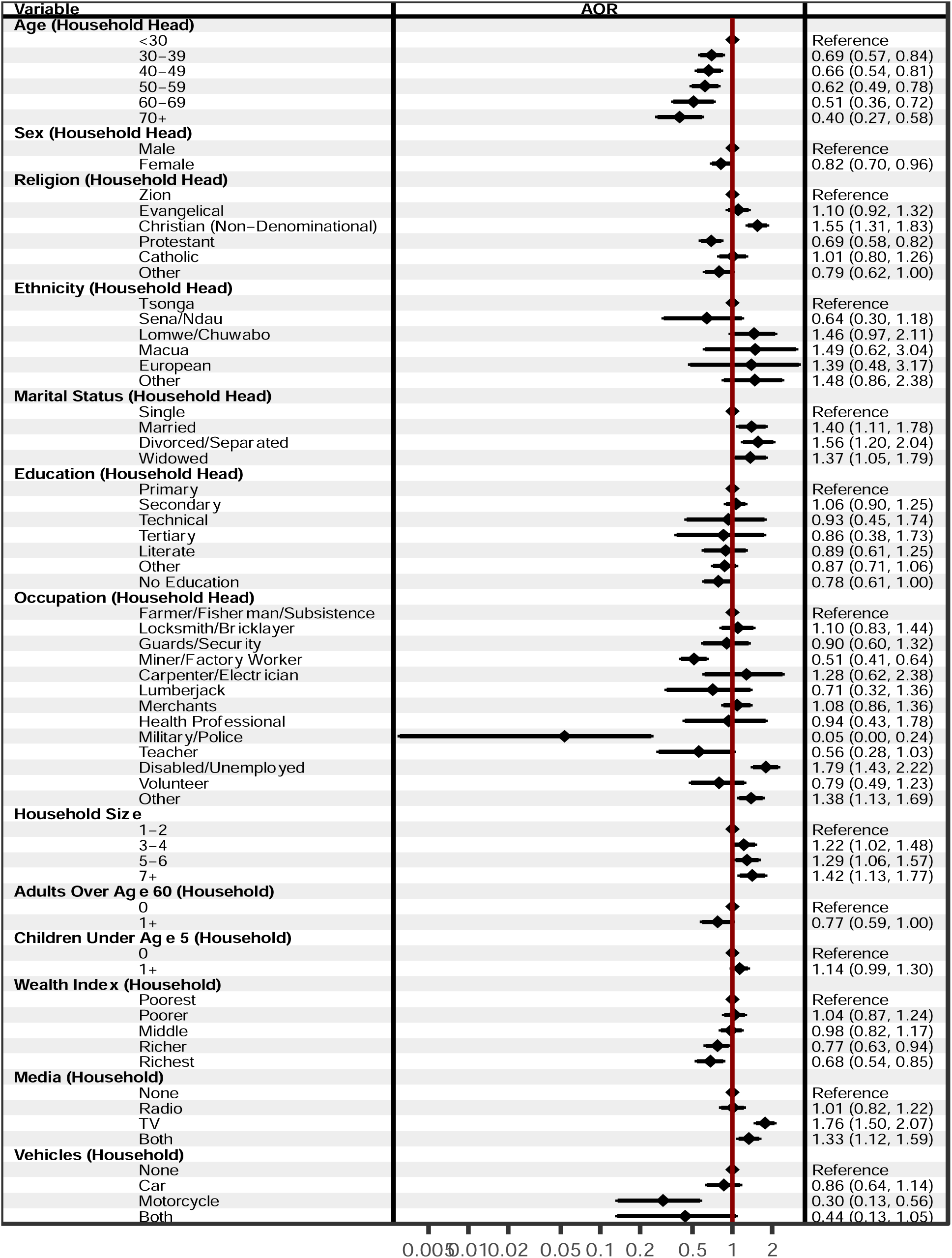
Adjusted Associations with Job Loss among Households Living in Manhiça (n=27,404). Presented as Adjusted Odds Ratios with 95% confidence intervals. Missing values for age, sex, religion, ethnicity, education, and occupation were less than 5 percent of the total observations.

**Figure B.5.**
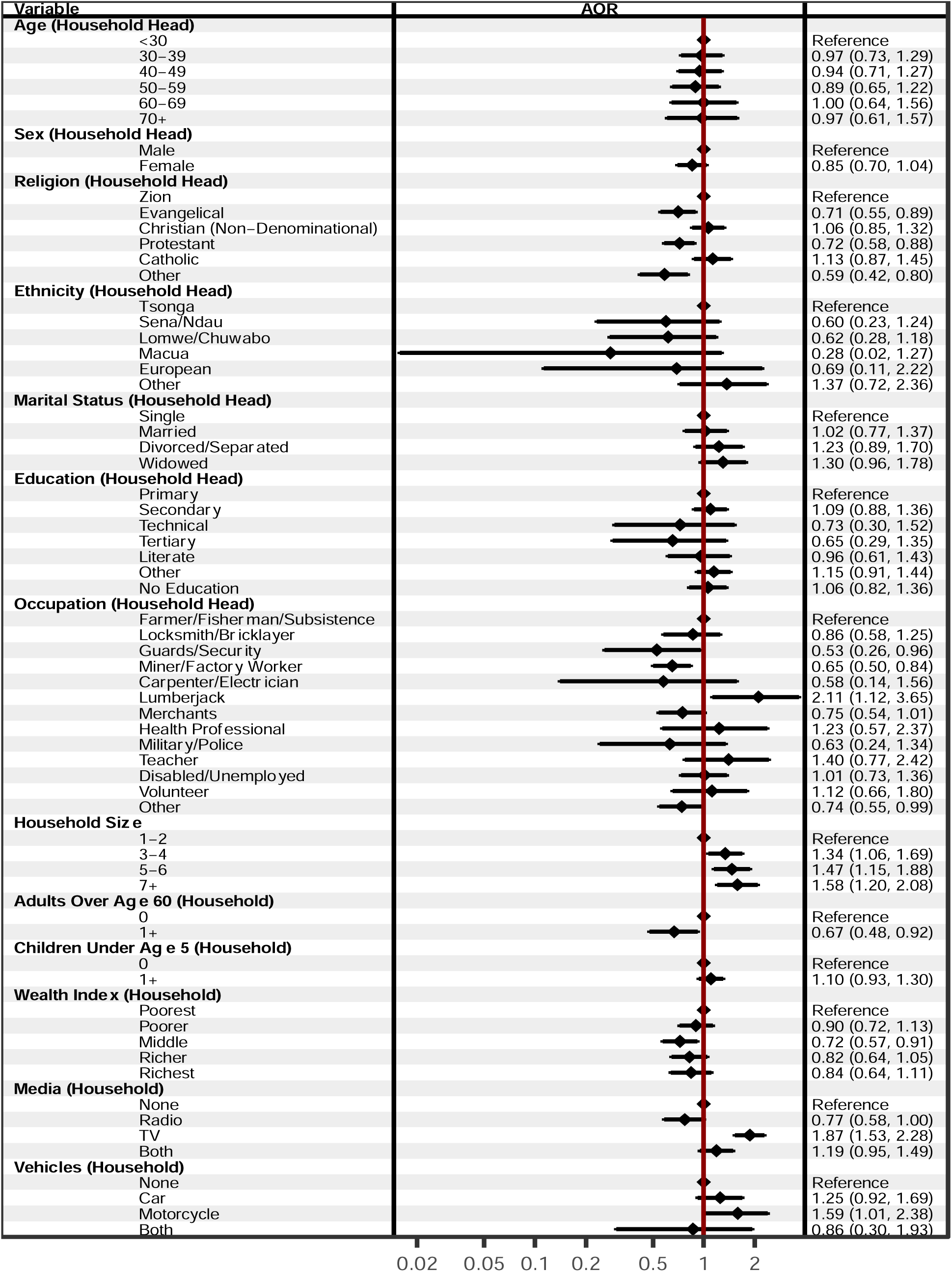
Adjusted Associations with Business Closures among Households Living in Manhiça (n=27,404). Presented as Adjusted Odds Ratios with 95% confidence intervals. Missing values for age, sex, religion, ethnicity, education, and occupation were less than 5 percent of the total observations.

## Footnotes

1 In preliminary analyses, these variables were used to generate additive and PCA based indices of household hardships; however, after evaluating these indices (e.g., the Cronbach’s Alpha scores for the additive index was 0.069) we concluded that the combination of these variables resulted in a poor fit for an index and therefore decided to examine each hardship independently.

